# Occupation and risk of severe COVID-19: prospective cohort study of 120,075 UK Biobank participants

**DOI:** 10.1101/2020.05.22.20109892

**Authors:** Miriam Mutambudzi, Claire L Niedzwiedz, Ewan B Macdonald, Alastair H Leyland, Frances S Mair, Jana J Anderson, Carlos A Celis-Morales, John G. Cleland, John Forbes, Jason MR Gill, Claire E Hastie, Frederick K Ho, Bhautesh D Jani, Daniel F Mackay, Barbara I Nicholl, Catherine A O’Donnell, Naveed Sattar, Paul Welsh, Jill P Pell, Srinivasa Vittal Katikireddi, Evangelia Demou

## Abstract

**Objectives:** To investigate severe COVID-19 risk by occupational group.

**Methods:** Baseline UK Biobank data (2006-10) for England were linked to SARS-CoV-2 test results from Public Health England (16 March to 26 July 2020). Included participants were employed or self-employed at baseline, alive and aged less than 65 years in 2020. Poisson regression models adjusted sequentially for baseline demographic, socioeconomic, work-related, health, and lifestyle-related risk factors to assess risk ratios (RRs) for testing positive in hospital or death due to COVID-19 by three occupational classification schemes (including Standard Occupation Classification 2000).

**Results:** Of 120,075 participants, 271 had severe COVID-19. Relative to non-essential workers, healthcare workers (RR 7.43, 95% CI:5.52,10.00), social and education workers (RR 1.84, 95% CI:1.21,2.82) and other essential workers (RR=1.60, 95% CI:1.05,2.45) had higher risk of severe COVID-19. Using more detailed groupings, medical support staff (RR 8.70, 95% CI:4.87,15.55), social care (RR 2.46, 95% CI:1.47,4.14) and transport workers (RR= 2.20, 95% CI:1.21,4.00) had highest risk within the broader groups. Compared to white non-essential workers, non-white non-essential workers had a higher risk (RR 3.27, 95% CI: 1.90,5.62) and non-white essential workers had the highest risk (RR 8.34, 95% CI:5.17,13.47). Using SOC2000 major groups, associate professional and technical occupations, personal service occupations and plant and machine operatives had higher risk, compared to managers and senior officials.

**Conclusions:** Essential workers have higher risk of severe COVID-19. These findings underscore the need for national and organizational policies and practices that protect and support workers with elevated risk of severe COVID-19.

****Trial registration**-N/A:**

What is already known on this topic
- Essential workers have a higher exposure to the SARS-CoV-2 virus due to the nature of their work.
- In comparison to non-essential workers, healthcare workers appear to have a higher risk of SARS-CoV-2 infection.

What this study adds
- Healthcare workers had a more than seven-fold higher risk of severe COVID-19; those working in social care and transport occupations had a two-fold higher risk.
- Adjusting for potential confounding and mediating variables did not fully account for the differences in the observed risk amongst most occupational groups.
- Non-white essential workers had the highest risk of severe COVID-19 infection.

How might this impact on policy or clinical practice in the foreseeable future?
- Our findings reinforce the need for adequate health and safety arrangements and provision of PPE, particularly in the health and social care sectors, and highlight the need for national and organizational policies and practices that protect and support workers with elevated risk of SARS-CoV-2 infection.

## INTRODUCTION

The Severe Acute Respiratory Syndrome coronavirus-2 (SARS-CoV-2) and its resulting disease (COVID-19) has resulted in a fast-moving pandemic. According to surveillance data from Public Health England (PHE) there were over 99,000 confirmed infections in England between January 31^st^, 2020 and April 22^nd^ 2020, with London reporting an incidence rate of 221/100,000 persons (1). Essential workers and older adults are particularly vulnerable to infection and adverse subsequent outcomes (2). At present however, few studies globally have assessed risk of COVID-19 in different essential worker groups and only one UK study has assessed COVID-19 related morbidity and mortality across different occupations, with limited consideration of potential confounding factors (3-6).

To protect public health, the UK instituted precautionary lockdown policies and urged businesses to transition to home working where possible during March 2020, to reduce risk of infection (7). However, the risks faced by different population groups during the shutdown have not been equal (8). Essential workers who provide crucial or fundamental public services including those in healthcare, social care, sanitary services, and transportation have continued attending work to carry out their daily duties. These essential worker groups have increased exposure to the SARS-CoV-2 virus through their work which may bring them into close proximity with members of the public or infected patients, particularly since carriers may be infectious without, or before, showing significant symptoms (6). In addition, their risk may be increased due to working closely with infected asymptomatic or even sick colleagues (presenteeism) who still report to work. Asymptomatic carriers and presenteeism in the workplace have both been associated with the spread of infectious diseases such as influenza and Ebola (9,10). Preliminary research indicates that occupational exposure to the SARS-CoV-2 virus is of great concern among essential worker groups, particularly healthcare workers, in whom the lack of personal protective equipment (PPE) caused “a real and justified fear about personal safety”(11). Inadequate PPE and challenges in implementing timely and effective practices in care homes has resulted in significant outbreaks in these occupational settings (12). In education, the reluctance to reopen schools because of concern about risk of infection could exacerbate existing inequalities (13). Furthermore, there is evidence of high infection rates and subsequent morbidity and mortality among low skilled occupations, and social, transport, food, and sales and retail workers (2,3,14-16).

Despite large occupational differences being generally seen for health outcomes (17), there has been a lack of studies examining differences in risk of COVID-19 across occupational groups. Apart from healthcare workers (18), it is not clear which other occupational groups are most at risk. Increasing our knowledge of the risk of infection among different groups of essential and non-essential workers will contribute to providing a more comprehensive depiction of the impact of global pandemics on vulnerable workers. This information has important implications for ensuring the safety and protection of essential workers from the risks of COVID-19 (19).

We therefore aimed to assess the risk of severe COVID-19 in essential workers, relative to nonessential workers. Specifically, we used linked data from the UK Biobank study and SARS-CoV-2 test results from Public Health England (PHE) to examine the risk of infection by a) broad essential occupational groups, b) detailed essential occupational groups and c) Standard Occupational Classification (SOC) 2000 major groups (20), while accounting for baseline sociodemographic, socioeconomic, work-related, lifestyle, and health factors.

## METHODS AND DATA

### Study design

UK Biobank is a prospective cohort study, established to identify disease determinants in middle and older age adults and has been previously described in detail (21). In brief, adults aged 40-69 years were invited to participate in the study if they resided within 25 miles (40.23 km) of an assessment centre and were registered with the National Health Service in England, Wales, or Scotland (22). Approximately 502,000 individuals (out of 9 million invited) consented to participate, representing a 5.5% response rate (21). At baseline participants were required to visit an assessment centre to complete a computer-assisted self-administered questionnaire and a face-to-face interview, and to provide physical measures and biological samples. All baseline data were collected between 2006 to 2010. The UK Biobank study received ethical approval from the NHS National Research Ethics Service North West (16/NW/0274) and all participants provided written informed consent.

UK Biobank participants who were: 1) working at baseline; 2) below retirement age (<65years) in 2020; 3) had their baseline assessment in England were included in the study. The latter criterion was used because linked SARS-CoV-2 test results from PHE were available for England only. Participants were excluded if they had previously requested to withdraw from the study (N=30).

### Ascertainment of outcomes

The outcome of interest was severe COVID-19, defined by a positive test result for SARS-CoV-2 in a hospital setting (i.e. participants whose tests were taken while an inpatient or attending an Emergency Department) or death with a primary or contributory cause reported as COVID-19 (International Classification of Disease-10 codes U07.1 or U07.2) (23). By focusing on hospital cases and deaths we limit potential bias due to differential ascertainment, as these cases likely reflect more severe COVID-19 disease and exclude those who were tested because they were a healthcare worker (1). Participants testing negative or positive outside a hospital setting were included in the denominator. We were not able to identify asymptomatic or symptomatic cases who did not present to the health service, therefore these were also included in the denominator.

Public Health England provided data for SARS-CoV-2 test results for the period 16 March 2020 to 26^th^ July 2020 from its microbiology database, Second Generation Surveillance System. Data provided included specimen date, origin (evidence that the individual was an inpatient or not) and result (positive or negative) (1). These data were linked to the UK Biobank baseline data and to mortality records from the NHS Information Centre up to 28^th^ June 2020.

### Ascertainment of exposure

Our exposure of interest was occupational group as reported at baseline. UK Biobank asked participants about their current or most recent job title and these were converted to 4 digit Standard Occupational Classification (SOC) 2000 codes (20). Employed participants were classified into five broad groups (non-essential workers, healthcare workers, social and education workers, police and protective service and ‘other’ essential workers) by members of our team with expertise in occupational and public health. To assess whether there were differences in risk among occupations within these broad groups, we further classified occupations into eight more precise categories of essential workers [healthcare professionals (e.g. doctors, pharmacists), health associate professionals (e.g. nurses, paramedics), medical support staff (nursing assistants, hospital porters), social care workers, education workers, food workers, transport workers, and police and protective services (including sanitary service workers)], whose risk was assessed relative to non-essential workers (see supplementary material Figure S1). Occupational groupings were performed blind to COVID-19 status.

To allow for comparability with research that uses occupations as defined by broader SOC groups, we also examined the associations between risk of severe COVID-19 and the SOC 2000 major occupation groups (managers and senior officials, professional occupations, associate professional and technical occupations, administrative and secretarial occupations, skilled trades occupations, personal service occupations, sales and customer service occupations, process, plant and machine operatives, elementary occupations) (5,20). As occupation data were collected at baseline between 2006-2010, we assessed correlations between occupation at baseline and follow-up for a subsample of the cohort (n=12,306) who participated in further data collection when attending a clinic visit to participate in the UK Biobank Imaging Study (24) between 30th April 2014 and 7th March 2019 (median August 2017). We found a high correlation (r=0.71, p<0.001) between job at baseline and follow-up indicating a high likelihood that participants had continued working in the same profession.

### Ascertainment of covariates

Covariates of interest included sociodemographic factors [current age group (<55, 55-59, 60+ years), gender (male/female), country of birth (UK and Ireland or elsewhere), ethnicity (white British, white Irish, white other, mixed, south Asian, black, other)], socioeconomic factors [area-level socioeconomic deprivation index, education level (college or university degree, A levels/AS levels or equivalent, O levels/GCSEs/CSEs or equivalent, other, none of the above)], work-related factors [shift work (yes, no), manual work (manual, non-manual), work hours (<40, 40-45, >45), tenure in job (<=10, 11-20, >20 years)], health conditions [number of self-reported chronic conditions, limiting illness/disability (yes, no)], and lifestyle-related factors [(alcohol consumption (daily or almost daily, three or four times a week, once or twice a week, one to three times a month, special occasions only, former drinker, never), smoking status (never, former, current), body mass index (BMI) category]. The Townsend index was used to assess area-level socioeconomic deprivation, which includes measures of neighborhood unemployment, non-car ownership, non-home ownership and household overcrowding (24). The index was categorised into quartiles reflecting a gradient from most advantaged (lowest quartile) to least advantaged (highest quartile). Self-reported number of chronic health conditions was ascertained from a pre-defined list of 43 conditions and categorized into none, one, two, three, four or more (25). BMI was calculated from physical measurements and treated as an ordinal variable with four categories according to the WHO classification (26): underweight (<18.5 kg/m^2^), normal (18.5-24.9 kg/m^2^), overweight (25.0-29.9 kg/m^2^), and obese (>30.0kg/m^2^). Assessment centre was included as a covariate in all models to account for potential differences in recruitment and measurement processes. All covariates were measured at baseline.

### Statistical analyses

Sample characteristics were summarised using frequencies and proportions. Poisson regression models for which risk ratios (RR) and 95% confidence intervals (95% CI) were reported, examined the strength of association between baseline occupational group and risk of severe COVID-19. Robust standard errors were used to ensure accurate estimation of 95% CIs and p values (27).

We estimated six nested models. Model 1 included sociodemographic factors, i.e. age, sex, assessment centre, country of birth, and ethnicity. Model 2 included all covariates in Model 1, plus socioeconomic factors, i.e. area-level socioeconomic deprivation quartile, and education level. Model 3 included all covariates in Model 2, plus work-related factors, i.e. shift work, manual work, job tenure, and work hours. Model 4 included all covariates in Model 2, plus number of chronic conditions, and longstanding illness/disability. Model 5 included the covariates from model 2 as well as lifestyle-related factors i.e. BMI, smoking, and alcohol. Model 6 was fully adjusted for all covariates. In post hoc analyses to examine potential effect modification by race, we grouped people into white/non-essential worker; nonwhite/non-essential worker; white/essential worker; non-white/essential worker and repeated the models above. Due to the small number of severe COVID-19 cases within groups when broken down by ethnicity, we were unable to investigate more detailed categories.

Participants with missing data (N=8,494 (6.6%)) for any variable were excluded from the statistical analyses. All analyses were performed using Stata MP/15.1 Software (Stata, College Station, TX).

### Patient and public involvement

Participants were not involved in the design and implementation of the study or in setting research questions and the outcome measures. No participants were asked to advise on interpretation or writing up of results.

## RESULTS

Our sample included 120,075 working participants aged 49 to 64 years in 2020, after excluding participants who died prior to 16^th^ March 2020 (n=2,067) and those with missing data (figure 1). Of these, 29.3% (n=35,127) were classified as essential workers; healthcare (9.0%), social and education (11.2%), and other essential workers (9.1 %) (Table 1). 92.2% of the sample was white (British, Irish, and other). South Asian and black participants accounted for 2.6%, and 2.7% of the study sample, respectively. Women and ethnic minority participants were more likely to be employed in essential occupations at baseline (supplementary table S1).

**Figure 1.**
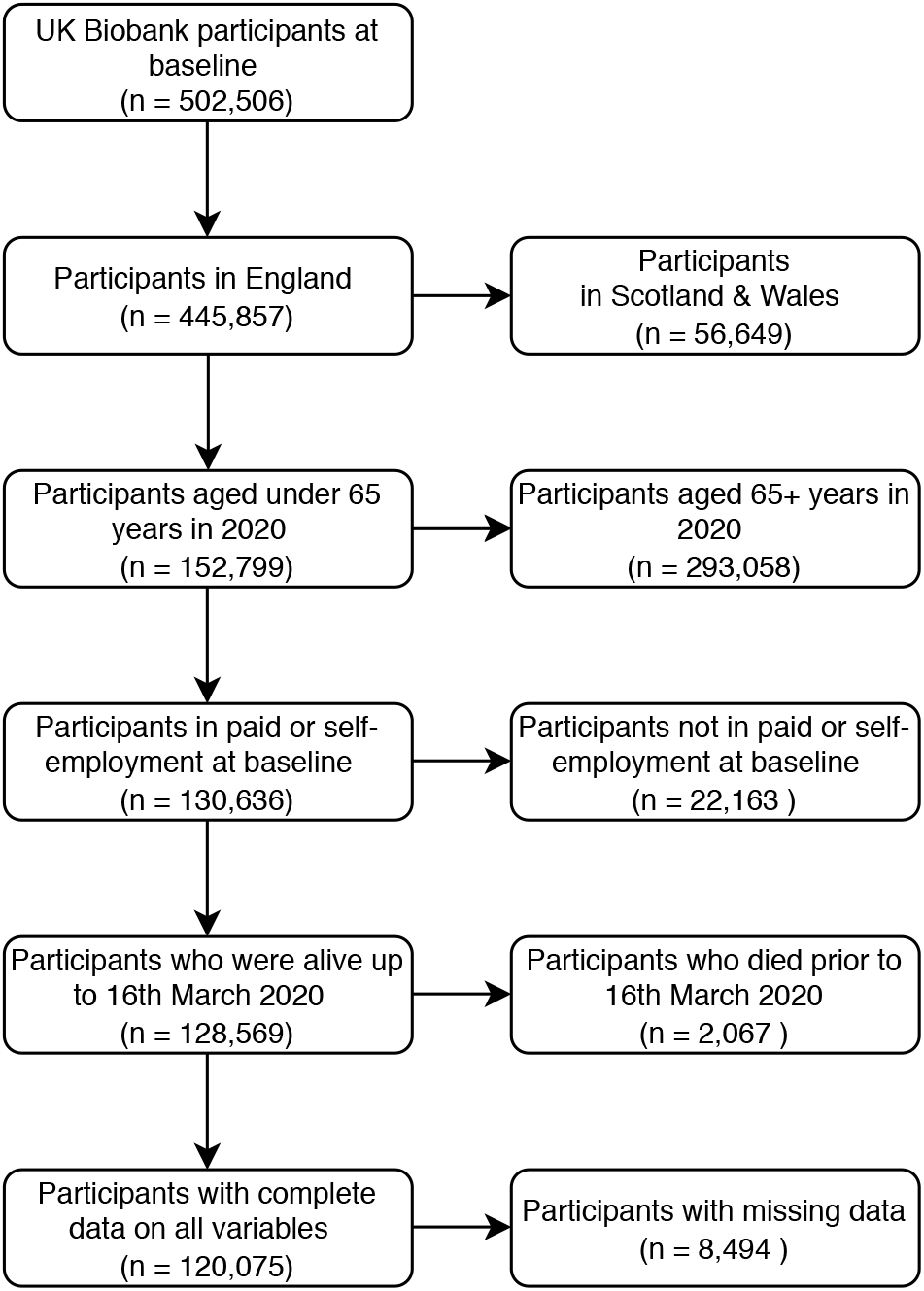
Flow chart of cohort

**Table 1:** Cohort characteristics for the sample of 120,075 UK Biobank participants recruited in 2006-10 and alive up to 16 March 2020

|  | N | % |
| --- | --- | --- |
| **Broad occupational groups of essential workers** |  |  |
| Non-essential workers | 84,948 | 70.7 |
| Healthcare workers | 10,748 | 9.0 |
| Social and education workers | 13,476 | 11.2 |
| Other essential workers | 10,903 | 9.1 |
| **Detailed occupational groups of essential workers** |  |  |
| Non-essential workers | 84,948 | 70.7 |
| Healthcare professionals | 1,779 | 1.5 |
| Medical support staff | 1,295 | 1.1 |
| Health associate professionals | 7,674 | 6.4 |
| Social care workers | 5,297 | 4.4 |
| Education workers | 8,179 | 6.8 |
| Food workers | 4,499 | 3.7 |
| Transport workers | 3,279 | 2.7 |
| Police and protective service workers | 3,125 | 2.6 |
| **SOC 2000 major occupational groups** |  |  |
| Managers and Senior Officials | 23,704 | 19.7 |
| Professional Occupations | 25,924 | 21.6 |
| Associate Professional and Technical Occupations | 23,054 | 19.2 |
| Administrative and Secretarial Occupations | 17,472 | 14.6 |
| Skilled Trades Occupations | 8,360 | 7.0 |
| Personal Service Occupations | 7,660 | 6.4 |
| Sales and Customer Service Occupations | 3,684 | 3.1 |
| Process, Plant and Machine Operatives | 4,792 | 4.0 |
| Elementary Occupations | 5,425 | 4.5 |
| **Age group (current)** |  |  |
| Under 55 | 25,315 | 21.1 |
| 55-59 | 44,734 | 37.3 |
| 60+ | 50,026 | 41.7 |
| **Sex** |  |  |
| Female | 65,063 | 54.2 |
| Male | 55,012 | 45.8 |
| **Ethnicity** |  |  |
| White British | 102,485 | 85.4 |
| White Irish | 3,205 | 2.7 |
| White Other | 4,974 | 4.1 |
| Mixed | 1,218 | 1.0 |
| South Asian | 3,075 | 2.6 |
| Black | 3,268 | 2.7 |
| Other | 1,850 | 1.5 |
| **Country of birth** |  |  |
| UK & Ireland | 108,159 | 90.1 |
| Elsewhere | 11,916 | 9.9 |
| **Education level** |  |  |
| College or University degree | 48,189 | 40.1 |
| A levels/AS levels or equivalent | 16,629 | 13.8 |
| O levels/GCSEs/CSEs or equivalent | 39,730 | 33.1 |
| Other | 10,157 | 8.5 |
| None of the above | 5,370 | 4.5 |
| **Deprivation quartile** |  |  |
| Quartile 1 (most advantaged) | 28,488 | 23.7 |
| Quartile 2 | 28,626 | 23.8 |
| Quartile 3 | 31,802 | 26.5 |
| Quartile 4 (least advantaged) | 31,159 | 25.9 |
| **Shiftwork** |  |  |
| No | 107,072 | 89.2 |
| Yes | 13,003 | 10.8 |
| **Manual occupation** |  |  |
| Non-manual | 103,634 | 86.3 |
| Manual | 16,441 | 13.7 |
| **Job tenure** |  |  |
| <=10 | 70,896 | 59.0 |
| 11-20 | 27,552 | 22.9 |
| >20 | 21,627 | 18.0 |
| **Working hours** |  |  |
| <40 | 61,946 | 51.6 |
| 40-45 | 38,279 | 31.9 |
| >45 | 19,850 | 16.5 |
| **Number of chronic conditions** |  |  |
| 0 | 61,244 | 51.0 |
| 1 | 38,526 | 32.1 |
| 2 | 14,374 | 12.0 |
| 3 | 4,319 | 3.6 |
| 4+ | 1,612 | 1.3 |
| **Long-standing illness, disability or infirmity** |  |  |
| No | 94,410 | 78.6 |
| Yes | 25,665 | 21.4 |
| **BMI Category** |  |  |
| Underweight (<18.5) | 613 | 0.5 |
| Normal weight (18.5-24.9) | 44,496 | 37.1 |
| Overweight (25.0-29.9) | 48,753 | 40.6 |
| Obese (≥30.0) | 26,213 | 21.8 |
| **Smoking status** |  |  |
| Never | 74,386 | 61.9 |
| Previous | 31,684 | 26.4 |
| Current | 14,005 | 11.7 |
| **Alcohol consumption** |  |  |
| Daily or almost daily | 20,080 | 16.7 |
| Three or four times a week | 29,942 | 24.9 |
| Once or twice a week | 35,273 | 29.4 |
| One to three times a month | 15,779 | 13.1 |
| Special occasions only | 11,985 | 10.0 |
| Never (former drinker) | 2,979 | 2.5 |
| Never | 4,037 | 3.4 |
| **Total** | 120,075 | 100.0 |

3,111 (2.6%) participants had been tested for SARS-CoV-2 between 16^th^ March and 26^th^ July 2020 and of these, 262 (0.2%) had a positive test in a hospital setting. Of the 262 hospital cases, 12 had died up to 28th June 2020 and an additional 9 people had COVID-19 as a contributory cause of death who were not identified as testing positive in hospital. 271 people (0.2%) were therefore classified as having severe COVID-19. Healthcare professionals (1.0%), medical staff support (1.1%), health associate professionals (0.9%), social care (0.3%), transport (0.4%) had higher rates of severe COVID-19 compared to nonessential workers (0.1%) (table 2). Descriptive statistics by broad race groups are included in supplementary table S2.

**Table 2:** Descriptive statistics for severe COVID-19 by occupational groups

|  | Severe COVID-19 |  |  |  |  |  |
| --- | --- | --- | --- | --- | --- | --- |
|  | No |  | Yes |  | Total | Total |
|  | N | % | N | % | N | % |
| **Broad occupational groups of essential workers** |  |  |  |  |  |  |
| Non-essential workers | 84,836 | 99.9 | 112 | 0.1 | 84,948 | 100 |
| Healthcare workers | 10,646 | 99.1 | 102 | 0.9 | 10,748 | 100 |
| Social and education workers | 13,445 | 99.8 | 31 | 0.2 | 13,476 | 100 |
| Other essential workers | 10,877 | 99.8 | 26 | 0.2 | 10,903 | 100 |
| **Detailed occupational groups of essential workers** |  |  |  |  |  |  |
| Non-essential workers | 84,836 | 99.9 | 112 | 0.1 | 84,948 | 100 |
| Healthcare professionals | 1,762 | 99.0 | 17 | 1.0 | 1,779 | 100 |
| Medical support staff | 1,281 | 98.9 | 14 | 1.1 | 1,295 | 100 |
| Health associate professionals | 7,603 | 99.1 | 71 | 0.9 | 7,674 | 100 |
| Social care workers | 5,279 | 99.7 | 18 | 0.3 | 5,297 | 100 |
| Education workers | 8,166 | 99.8 | 13 | 0.2 | 8,179 | 100 |
| Food workers | 4,492 | 99.8 | 7 | 0.2 | 4,499 | 100 |
| Transport workers | 3,267 | 99.6 | 12 | 0.4 | 3,279 | 100 |
| Police and protective service workers | 3,118 | 99.8 | 7 | 0.2 | 3,125 | 100 |
| **SOC 2000 major occupational groups** |  |  |  |  |  |  |
| Managers and Senior Officials | 23,675 | 99.9 | 29 | 0.1 | 23,704 | 100 |
| Professional Occupations | 25,879 | 99.8 | 45 | 0.2 | 25,924 | 100 |
| Associate Professional and Technical Occupations | 22,960 | 99.6 | 94 | 0.4 | 23,054 | 100 |
| Administrative and Secretarial Occupations | 17,444 | 99.8 | 28 | 0.2 | 17,472 | 100 |
| Skilled Trades Occupations | 8,351 | 99.9 | 9 | 0.1 | 8,360 | 100 |
| Personal Service Occupations | 7,632 | 99.6 | 28 | 0.4 | 7,660 | 100 |
| Sales and Customer Service Occupations | 3,677 | 99.8 | 7 | 0.2 | 3,684 | 100 |
| Process, Plant and Machine Operatives | 4,775 | 99.6 | 17 | 0.4 | 4,792 | 100 |
| Elementary Occupations | 5,411 | 99.7 | 14 | 0.3 | 5,425 | 100 |
| Total | 119,804 | 99.8 | 271 | 0.2 | 120,075 | 100 |

### Risk of severe COVID-19 by broad essential occupational groups

In comparison to non-essential workers, healthcare workers had a more than seven-fold (RR 7.43, 95% CI: 5.52,10.00) greater risk of severe COVID-19 (table 3). This association remained after adjusting for all covariates (RR 7.69, 95% CI: 5.58,10.60). Social and education workers also exhibited a higher risk (RR 1.84, 95% CI: 1.21,2.82), which remained after adjustment for the covariates. Other essential workers also had slightly higher risk compared to non-essential workers (RR 1.60, 95% CI: 1.05,2.45), but this was attenuated after adjustment for socioeconomic factors. Detailed model results including all covariates are presented in supplementary table S3. In summary, men, south Asian and black ethnic groups, socioeconomic disadvantage and the least educated groups had higher risk of severe COVID-19, compared to women, white British, socioeconomic advantage and degree educated groups, respectively. Work-related factors including shift-work and manual work were also associated with higher risk of severe COVID-19, as were being overweight or obese, or a previous smoker.

**Table 3:** Risk ratios for severe COVID-19 by occupational groups (n=120,075)

|  | **Model 1** | **Model 2** | **Model 3** | **Model 4** | **Model 5** | **Model 6** |
| --- | --- | --- | --- | --- | --- | --- |
| **Severe COVID-19** | **RR** | **RR** | **RR** | **RR** | **RR** | **RR** |
|  | **[95% CI]** | **[95% CI]** | **[95% CI]** | **[95% CI]** | **[95% CI]** | **[95% CI]** |
| **Broad occupational groups of essential workers** |  |  |  |  |  |  |
| Non-essential workers (reference) | 1 | 1 | 1 | 1 | 1 | 1 |
| Healthcare workers | 7.43***<br>[5.52,10.00] | 8.45***<br>[6.22,11.47] | 7.57***<br>[5.50,10.41] | 8.44***<br>[6.21,11.46] | 8.53***<br>[6.29,11.58] | 7.69***<br>[5.58,10.60] |
| Social and education workers | 1.84**<br>[1.21,2.82] | 2.00**<br>[1.30,3.08] | 1.90**<br>[1.22,2.94] | 1.99**<br>[1.29,3.06] | 1.97**<br>[1.28,3.03] | 1.88**<br>[1.21,2.91] |
| Other essential workers | 1.60*<br>[1.05,2.45] | 1.30<br>[0.85,1.98] | 1.17<br>[0.76,1.80] | 1.30<br>[0.85,1.98] | 1.27<br>[0.83,1.94] | 1.15<br>[0.75,1.77] |
| **Detailed occupational groups of essential workers** |  |  |  |  |  |  |
| Non-essential workers (reference) | 1 | 1 | 1 | 1 | 1 | 1 |
| Healthcare professionals | 6.19***<br>[3.68,10.43] | 8.62***<br>[4.98,14.94] | 8.26***<br>[4.77,14.28] | 8.70***<br>[5.02,15.06] | 9.33***<br>[5.40,16.14] | 8.99***<br>[5.20,15.54] |
| Medical support staff | 8.70*** | 7.43*** | 6.48*** | 7.39*** | 7.33*** | 6.42*** |
|  | [4.87,15.55] | [4.17,13.25] | [3.62,11.58] | [4.13,13.19] | [4.13,13.02] | [3.60,11.45] |
| Health associate professionals | 7.53***<br>[5.44,10.43] | 8.54***<br>[6.13,11.90] | 7.61***<br>[5.33,10.87] | 8.52***<br>[6.11,11.88] | 8.54***<br>[6.12,11.92] | 7.65***<br>[5.34,10.95] |
| Social care workers | 2.46***<br>[1.47,4.14] | 2.38**<br>[1.42,4.00] | 2.19**<br>[1.29,3.72] | 2.36**<br>[1.40,3.97] | 2.31**<br>[1.37,3.88] | 2.13**<br>[1.25,3.63] |
| Education workers | 1.36<br>[0.75,2.48] | 1.61<br>[0.88,2.96] | 1.59<br>[0.86,2.92] | 1.61<br>[0.88,2.95] | 1.62<br>[0.88,2.96] | 1.59<br>[0.87,2.91] |
| Food workers | 1.12<br>[0.52,2.42] | 0.93<br>[0.43,1.98] | 0.85<br>[0.40,1.83] | 0.93<br>[0.43,1.98] | 0.92<br>[0.43,1.96] | 0.84<br>[0.39,1.80] |
| Transport workers | 2.20**<br>[1.21,4.00] | 1.66<br>[0.91,3.01] | 1.48<br>[0.81,2.70] | 1.66<br>[0.91,3.01] | 1.58<br>[0.87,2.90] | 1.43<br>[0.78,2.63] |
| Police and protective service workers | 1.55<br>[0.72,3.32] | 1.36<br>[0.63,2.93] | 1.21<br>[0.56,2.64] | 1.35<br>[0.62,2.92] | 1.32<br>[0.61,2.86] | 1.19<br>[0.55,2.58] |
| **SOC 2000 major occupational groups** |  |  |  |  |  |  |
| Managers and Senior Officials (reference) | 1 | 1 | 1 | 1 | 1 | 1 |
| Professional Occupations | 1.36<br>[0.85,2.18] | 1.47<br>[0.91,2.36] | 1.49<br>[0.92,2.41] | 1.47<br>[0.91,2.36] | 1.51<br>[0.94,2.43] | 1.53<br>[0.95,2.48] |
| Associate Professional and Technical Occupations | 3.19***<br>[2.10,4.85] | 3.11***<br>[2.05,4.72] | 2.73***<br>[1.77,4.23] | 3.10***<br>[2.04,4.71] | 3.15***<br>[2.08,4.79] | 2.78***<br>[1.79,4.29] |
| Administrative and Secretarial Occupations | 1.24<br>[0.73,2.12] | 1.14<br>[0.67,1.95] | 1.22<br>[0.71,2.11] | 1.14<br>[0.67,1.94] | 1.17<br>[0.68,2.00] | 1.24<br>[0.72,2.15] |
| Skilled Trades Occupations | 0.82<br>[0.39,1.74] | 0.67<br>[0.31,1.44] | 0.49<br>[0.22,1.06] | 0.68<br>[0.32,1.45] | 0.69<br>[0.32,1.49] | 0.50<br>[0.23,1.09] |
| Personal Service Occupations | 2.73***<br>[1.56,4.76] | 2.31**<br>[1.32,4.03] | 1.75<br>[0.99,3.10] | 2.29**<br>[1.32,4.00] | 2.33**<br>[1.34,4.06] | 1.77<br>[1.00,3.13] |
| Sales and Customer Service Occupations | 1.36<br>[0.59,3.17] | 1.09<br>[0.46,2.57] | 0.91<br>[0.38,2.18] | 1.08<br>[0.46,2.55] | 1.08<br>[0.46,2.56] | 0.90<br>[0.38,2.16] |
| Process, Plant and Machine Operatives | 2.39**<br>[1.31,4.36] | 1.82<br>[0.99,3.34] | 1.25<br>[0.66,2.36] | 1.81<br>[0.99,3.33] | 1.81<br>[0.99,3.34] | 1.26<br>[0.67,2.37] |
| Elementary Occupations | 1.76<br>[0.92,3.34] | 1.29<br>[0.65,2.53] | 0.87<br>[0.43,1.75] | 1.28<br>[0.65,2.52] | 1.31<br>[0.67,2.59] | 0.89<br>[0.44,1.79] |
| **Observations** | 120075 | 120075 | 120075 | 120075 | 120075 | 120075 |

CI=confidence interval; RR=risk ratio
\* $p < 0.05$ , \*\* $p < 0.01$ , \*\*\* $p < 0.001$
Coefficients for the covariates not shown.
Model 1: Adjusted for age group, sex, ethnicity, country of birth
Model 2: Model 1 + socioeconomic deprivation quartile, education level
Model 3: Model 2 + shift work, manual work, job tenure, working hours
Model 4: Model 2 + number of chronic conditions, long-standing illness/disability
Model 5: Model 2 + BMI category, smoking status, alcohol consumption
Model 6: All covariates

### Risk of severe COVID-19 by detailed essential occupational groups

Examination of associations using more detailed occupation profiles (figure 2a) indicated that relative to non-essential workers, medical support staff had the highest risk of severe COVID-19 (RR 8.70, 95% CI: 4.87,15.55), followed by health associate professionals (RR 7.53, 95% CI: 5.44,10.43) and healthcare professionals (RR 6.19, 95% CI: 3.68,10.43) (table 3). The higher risk of severe COVID-19 among healthcare workers was not reduced after adjustment for socioeconomic, work-related, or health and lifestyle-related factors. Among social care workers, risk was also elevated (RR 2.46, 95% CI: 1.47,4.14) and was only slightly attenuated when adjusting for the covariates. Transport workers also exhibited a two-fold higher risk of severe COVID-19 (RR 2.20, 95% CI: 1.21,4.00) compared to non-essential workers, but this was attenuated after adjustment for socioeconomic factors (RR 1.66, 95% CI: 0.91,3.01). There were no strong associations observed for the other essential worker groups (police and protective service, food, or education workers). Further details for these models are presented in supplementary table S4.

**Figure 2:**
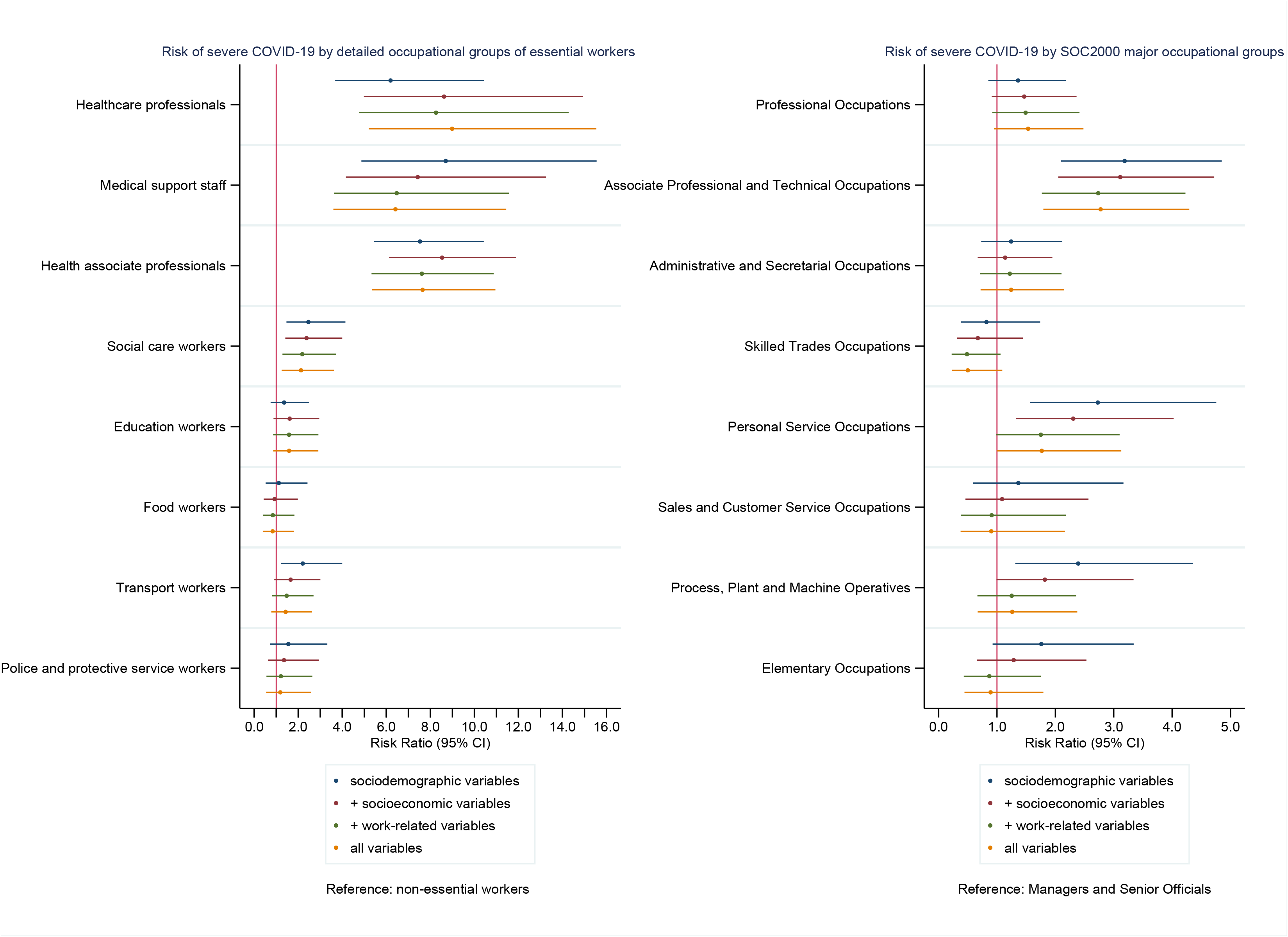
Risk ratios for the associations between (a) detailed essential occupational groups, (b) SOC2000 major occupational groups and severe COVID-19

### Risk of severe COVID-19 by SOC 2000 major occupational groups

In analyses using the SOC 2000 major occupational groups (table 3 and figure 2b), compared to managers and senior officials, associate professional and technical occupations (RR 3.19, 95% CI: 2.10,4.85) had the highest risk, which was only slightly attenuated by adjusting for covariates. Personal service occupations were associated with higher risk (RR 2.73, 95% CI: 1.56,4.76), but this was attenuated after adjustment for the covariates, particularly work-related factors including shift and manual work. Process, plant and machine operatives (RR 2.39, 95% CI: 1.31,4.36) also had a higher risk, however this was mostly explained by socioeconomic factors. The other occupational groups (professional, administrative and secretarial, skilled trades, sales and customer service and elementary occupations) did not have elevated risk. Detailed model results for the association between SOC 2000 major occupational groups and severe COVID-19 are available in supplementary table S5.

### Post hoc analyses

In post hoc analyses examining potential effect modification by race, we found that the risk of severe COVID-19 was highest in non-white, essential workers, with a more than 8-fold risk (RR 8.34, 95% CI: 5.17,13.47) compared to non-essential workers who were white (table S6 and figure S2). The risks for nonwhite, non-essential workers (RR 3.27, 95% CI: 1.90,5.62) and white, essential workers (RR 3.47, 95% CI 2.63,4.59) were similar, suggesting effect modification by race. Accounting for the range of socioeconomic, health, work and lifestyle-related factors did not substantially attenuate the associations.

## DISCUSSION

To our knowledge, our study is the largest to date to assess risk of severe COVID-19 among different occupational groups. We found an over seven-fold higher risk for healthcare workers, and a twofold higher risk for social care and transport workers, compared to non-essential workers. Apart from transport workers, adjustment for covariates did not alter the associations substantially, implying that the socioeconomic, health, work- and lifestyle-related variables we studied were not the main mechanistic factors underpinning the occupational differences. The heightened risk found among transport workers appeared to be accounted for by socioeconomic factors. The comparisons of severe COVID-19 risk across health and social care occupational groups highlighted how these higher risks seem to be particularly linked to the jobs themselves, rather than reflecting broader socioeconomic circumstances.

This study has several important strengths. First, by using a well characterised cohort study, we were able to compare infection risk across a wide range of occupational groups and identify occupations that may be at higher risk of severe COVID-19. Data linkage, the large sample size and detailed data, enabled us to expeditiously provide empirical evidence from the ongoing pandemic and to investigate the extent to which observed outcomes are potentially explained by a wide range of factors, including work-related factors, lifestyle risk factors, pre-existing health status and other social variables.

Our findings should be considered in light of several limitations. Baseline data were collected 10-14 years ago, and we are unable to fully account for potential changes in health, lifestyle, sociodemographic and employment status. Further, the UK Biobank has low participation from ethnic minorities and low-income adults (28). As participation in research studies is non-random this may lead to collider bias and increase the risk of inaccurate associations not generalizable to the general population (29,30). The number of cases does not allow for an assessment of risk of COVID-19 for more detailed occupational groups and necessitates the grouping of occupations into broad exposure categories, which may have led to some exposure misclassification. Multiple testing may increase the probability of false positives, but using only our primary outcome of severe COVID-19 risk and broad subgroups mitigates this issue (31). Our results also reflect circumstances during the early phase of the pandemic in March to July 2020. Risks may differ over time, as the extent of physical distancing measures, work patterns and structures or availability of PPE changes. Our outcome measure is also a measure of severe acute disease and so results may be different for asymptomatic cases, those who experienced symptoms who were not tested, or those who experience long-term effects (32).

Our findings are corroborated by preliminary research reporting higher risk of COVID-19 in essential workers (2,14-16,18). Recent UK Office for National Statistics (ONS) data examining mortality from COVID-19 however, suggests a slightly different pattern of findings from those we present here (5). ONS reported high COVID-19 death rates in men in the lowest skilled occupations, but similarly find higher mortality rates among male healthcare workers, transport and social care workers (5). Several reasons may explain why they find higher risk among elementary occupations. The key reason is likely due to their inclusion of people aged 20 to 64 years, whereas our sample is mostly people aged 50 to 64 years and so is affected by survival bias. Low-skilled workers are disproportionately affected by socioeconomic disadvantage (33), which is associated with poorer health outcomes and higher mortality rates overall (17,34).

There is an urgent need for policies and workplace interventions to reduce exposure and limit spread of infectious diseases in the workplace, through ensuring availability of resources for protective equipment and training. Interventions should be rapidly implemented and delivered, based on the best available evidence, especially as other occupational groups return to workplaces and social distancing measures are relaxed (35). Combining our findings with those of the ONS (5), it is clear that maintaining testing for essential workers is important; however, there is an urgent need for testing and protective measures to be extended to wider and more disadvantaged occupational groups.

Future research will need to assess risk differences among other working groups, such as younger workers and monitor how the progression of COVID-19 and its long-term effects may impact different occupational groups. Ethnic (36,37) and occupational (3,5) inequalities in SARS-CoV-2 exposure, infection, and mortality are evident and these should be studied in combination. Unfortunately, our sample did not allow for detailed analysis, but in our post hoc analyses we found that non-white essential workers were disproportionally at higher risk of severe COVID-19. Our study findings reinforce the need for adequate health and safety arrangements and provision of PPE for essential workers especially in the health and social care sectors, which should be urgently addressed. The health and wellbeing of essential workers is critical to limiting the spread, and managing the burden of global pandemics (38).

## Data Availability

This research has been conducted using the UK Biobank Resource (application No 41686 & 17333).

## ACKNOWLEDGEMENTS

We thank the UK Biobank participants. This research has been conducted using the UK Biobank Resource (application No 41686 & 17333). We also acknowledge financial support from the Medical Research Council and Chief Scientist Office (MC_UU_12017/13; SPHSU13). CLN is supported by a Medical Research Council Fellowship (MR/R024774/1) and SVK by a NRS Senior Clinical Fellowship (SCAF/15). The views and opinions expressed are those of the authors and do not necessarily reflect those of the above funding bodies.

